# Multimodal Classification of Mild Traumatic Brain Injury Reveals Local Coupling Between Structural and Functional Connectomes

**DOI:** 10.1101/2021.12.14.21267815

**Authors:** Ajay Peddada, Kevin S. Holly, Tejaswi D. Sudhakar, Christina Ledbetter, Christopher E. Talbot, Daniel Valdivia, Piyush Kalakoti, Elizabeth Ginalis, Travis Quinoa, Benjamin J. Barker, Derrick Murcia, Rebekah Daggett, Phillip Holly, Tina Phan, Robert C. Ross, Eduardo Gonzalez-Toledo, Bharat Biswal, Hai Sun

## Abstract

**Background:** Following mild traumatic brain injury (mTBI) compromised white matter structural integrity can result in alterations in functional connectivity of large-scale brain networks and may manifest in functional deficit including cognitive dysfunction. Advanced magnetic resonance neuroimaging techniques, specifically diffusion tensor imaging (DTI) and resting state functional magnetic resonance imaging (rs-fMRI), have demonstrated an increased sensitivity for detecting microstructural changes associated with mTBI. Identification of novel imaging biomarkers can facilitate early detection of these changes for effective treatment. In this study, we hypothesize that feature selection combining both structural and functional connectivity increases classification accuracy.

**Methods:** 16 subjects with mTBI and 20 healthy controls underwent both DTI and resting state functional imaging. Structural connectivity matrices were generated from white matter tractography from DTI sequences. Functional connectivity was measured through pairwise correlations of rs-fMRI between brain regions. Features from both DTI and rs-fMRI were selected by identifying five brain regions with the largest group differences and were used to classify the generated functional and structural connectivity matrices, respectively. Classification was performed using linear support vector machines and validated with leave-one-out cross validation.

**Results:** Group comparisons revealed increased functional connectivity in the temporal lobe and cerebellum as well as decreased structural connectivity in the temporal lobe. After training on structural connections only, a maximum classification accuracy of 78% was achieved when structural connections were selected based on their corresponding functional connectivity group differences. After training on functional connections only, a maximum classification accuracy of 69% was achieved when functional connections were selected based on their structural connectivity group differences. After training on both structural and functional connections, a maximum classification accuracy of 69% was achieved when connections were selected based on their structural connectivity.

**Conclusions:** Our multimodal approach to ROI selection achieves at highest, a classification accuracy of 78%. Our results also implicate the temporal lobe in the pathophysiology of mTBI. Our findings suggest that white matter tractography can serve as a robust biomarker for mTBI when used in tandem with resting state functional connectivity.

## 1 Introduction

White matter fiber tracts constitute the structural pathways that interlink distinct brain regions, forming the anatomical backbone of structural connectivity networks (Wang, 2020). The integrity of white matter tracts is essential for normal brain function. These structural connectivity networks support functional interactions between brain areas and thus establish a structure-function relationship (Craddock et al. 2013). In mild traumatic brain injury (mTBI), traumatic insult to these structural networks results in axonal injury. As a result, compromised structural integrity can initiate compensatory changes in structural networks (Andriessen, 2010; Chatelin, 2011). This can cause collateral alterations in functional connectivity of large-scale brain networks. These abberant changes may manifest clinically with symptoms or impairments in cognitive, sensorimotor, or behavioral function (Harris, Verley, Gutman, Thompson, et al. 2016; Sinke et al. 2021). Therefore, mTBI results in a complex pattern of network dysfunction (Hayes, Bigler, and Verfaellie 2016; Mckee and Daneshvar 2015).

Advanced neuroimaging techniques have been shown to capture network-level dysfunction in mTBI. Diffusion tensor imaging (DTI) has demonstrated an increased sensitivity for detecting microstructural changes, such as DAI (Puig et al. 2020). DTI generates signal contrast when proton diffusion is anisotropic, appropriate for visualizing a highly organized fiber structure. Random and isotropic diffusion of protons, reflective of unrestricted water dispersion due to loss of white matter integrity, results in the loss of signal contrast (Harris, Verley, Gutman, and Sutton 2016; Hayes, Bigler, and Verfaellie 2016). Furthermore, diffusion tractography has been shown to adequately quantify structural connectivity. However, literature reports DTI tractography to both underestimate and overestimate white matter fiber connections by inadequately quantifying weak long-range connections as well as over quantifying spurious connections, increasing both false-negative and false-positive results, respectively (Chu, Parhi, and Lenglet 2018; R. E. Smith et al. 2012).

Network alterations following mTBI have also been investigated using functional magnetic resonance imaging (fMRI) and is well reported in the literature (Mayer et al. 2011; Stevens et al. 2012; Palacios et al. 2017; Iraji et al. 2015). Resting state functional magnetic resonance imaging (rs-fMRI) quantifies blood oxygen levels as a surrogate marker of brain activity, and functional connectivity measures the temporal correlation between different brain regions in resting state brain networks (Sharp et al. 2011; Biswal et al. 1995). rs-fMRI analysis of mTBI has characterized alterations in functional activity potentially due to direct injury of functional networks or remodeling following traumatic insult (Mayer et al. 2011).

Recent studies have shown that multimodal methods combining structural and functional information, quantified by DTI tractography and rs-fMRI, can better detect compromised network integrity (Chu, Parhi, and Lenglet 2018). Multiple studies have reported on the relationship between structural and functional connectivity in mild TBI (Harris, Verley, Gutman, and Sutton 2016; Harris, Verley, Gutman, Thompson, et al. 2016; Iraji et al. 2016a; Tang et al. 2012). A study by Sharp et al. in 2011 reported decreased functional connectivity in the default mode network (DMN) in mTBI subjects with decreased structural connectivity (Sharp et al. 2011). Palacios and colleagues reported an inverse relationship, exhibiting decreased structural connectivity and increased functional connectivity in regions in the frontal lobe in chronic traumatic brain injury subjects. (Palacios et al. 2013a).

The emergence of large, multimodal datasets has enabled the use of multivariate statistical modeling techniques known as “machine learning” to both predict pathologic conditions as well as extract potential biomarkers. Machine learning has been used on single imaging modalities such as DTI tractography (Mitra et al. 2016) has also achieved strong classification performance on larger datasets combining T1 weighted MRI and other advanced imaging sequences (Lui et al. 2014).

However, not all studies have found success in classification with multimodal imaging data. Vergara et al found that combining both structural and functional connectivity reduced classification performance (Vergara et al. 2017).

In our study, we quantified changes in functional and structural connectomes between mTBI and healthy controls. We showed that group differences in functional connectivity can help identify structural connections predictive of mTBI. To this end, we trained a multivariate machine learning algorithm to classify subjects with mTBI from healthy controls. We found that feature selection using multimodal imaging improved classification accuracy. Through this method, we were able to identify a set of brain regions that are particularly vulnerable to mTBI.

## 2 Methods

### 2.1 Subjects

A retrospective chart review was conducted to identify patients with mTBI who underwent neuroimaging at LSUHSC between September 2015 and June 2017. For the control cohort, subjects with matched acquisition parameters were identified from an in-house normal control database and utilized for this study. Approval for this study was granted by Louisiana State University Health Sciences Center (LSUHSC) Institutional Review Board (IRB). Brain images of thirty-three subjects with mTBI and thirty-four healthy controls were used in the study. After removing subjects whose scans were affected by artifacts, sixteen subjects with mTBI and twenty control subjects were included in this analysis. Age and sex of mTBI and control subjects were compared using independent two-tailed t-tests with the SciPy library in Python (Jones, Oliphant, and Peterson 2001; Rossum and Drake 1995).

### 2.2 Image Acquisition

In this study, all MRI scans were acquired on a single 1.5 T clinical MR systems (GE Medical Systems, Milwaukee, WI, USA). The MRI examination included a high resolution, non-contrast enhanced T1-weighted sequence (TR/TE 9.644/3.82, 90° flip angle, 256 × 256 matrix size, 1.2-mm slice thickness), diffusion tensor sequence (An optimized TE, 90° flip angle, 256 × 256 matrix size, field of view 28 cm, 5-mm slice thickness, 1 mm spacing, axial slice orientation, 36 directions, b-values, 1000; NEX, 1), and EPI-BOLD functional MRI sequence (TR/TE 3,000/60, 64 × 64 matrix size, 5-mm slice thickness, 5 min 12 sec, 104 whole brain resting state acquisition, where the first 4 were discarded). Retrieved DICOM images for eligible participants were converted to NIFTI format using MRIcron (www.mccauslandcenter.sc.edu/crnl/mricron/).

Using 3D Slicer version 4.1.1 (http://www.slicer.org), T1 scans were registered to the baseline DTI volume and saved in the NIfTI (.nii) format (See supplementary material for details) (Andriy et al. 2012). BrainSuite version 16a1 was used to generate brain masks from T1 sequences (www.brainsuite.org).

The brain masks were manually edited within BrainSuite by 4 individuals who checked each other’s work to ensure high quality.

### 2.3 fMRI Preprocessing

Preprocessing of fMRI scans was done using the default pipeline in the CONN-fMRI Functional Connectivity toolbox, which implements SPM12 (Whitfield-Gabrieli and Nieto-Castanon 2012; Friston 2007). The preprocessing steps were functional realignment and unwarping, slice-timing correction, outlier identification, direct segmentation and normalization, and functional smoothing. Data was transformed to the Montreal Neurological Institute standard space at a resolution of 2 × 2 × 2 mm^3^. Data was smoothed with an 8 mm full width at half maximum (FWHM) kernel. Images with a framewise displacement above 0.9mm or BOLD signal changes 5 standard deviations above the global mean were flagged as outliers.

### 2.4 Establishing Nodes for the Connectome

The Harvard-Oxford cortical atlas and AAL subcortical atlas was imported from CONN into BrainSuite, which contained Brodmann areas from the Talairach Daemon atlas (www.talairach.org) along with four Fox nodes that were brought into MNI-space through a Lancaster transform (Whitfield-Gabrieli and Nieto-Castanon 2012). In CONN, the global signal was regressed. This atlas defined 132 regions of interests (ROIs), i.e., nodes. MNI coordinates for all nodes are listed in Table S1. These nodes are used to estimate both structural and functional connectivity metrics. From these structural and functional connectivity metrics, connectivity matrices were then computed among the nodes.

### 2.5 Computing Functional Connectivity Matrix from rs-fMRI

T1 weighted and rs-fMRI sequences were preprocessed within the open-source software package, CONN, using Statistical Parametric Mapping software (SPM12, Wellcome Department of Imaging Neuroscience, Institute of Neurology and the National Hospital for Neurology and Neurosurgery; London, England). As shown in Figure 1, CONN was utilized to analyze FC based on the rs-fMRI sequences within nodes that were segmented based on the high-resolution T1 anatomical image (Whitfield-Gabrieli and Nieto-Castanon 2012). The FC strength between nodes was determined by a Fischer transform correlation coefficient within CONN, which measures the correlation of BOLD signals between the two nodes. Connectivity matrices containing these correlation coefficients for each node-to-node were exported from CONN.

**Figure 1.**
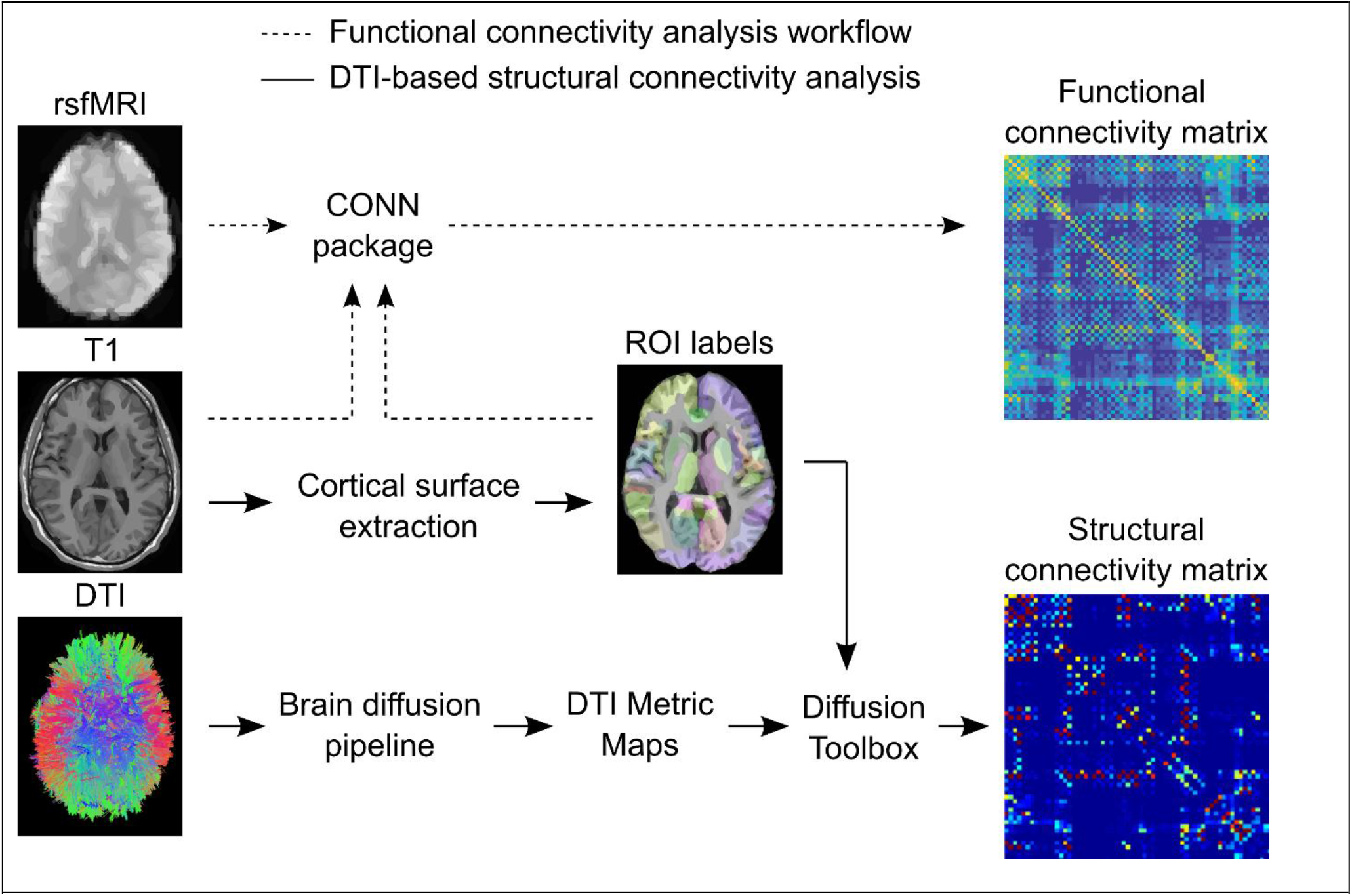
Workflows for generating functional and structural connectivity matrices.

### 2.6 Preprocessing DTI

Voxel-wise statistical analysis of the fractional anisotropy (FA) data was carried out using TBSS (Tract-Based Spatial Statistics, (S. M. Smith et al. 2006)), part of FSL (S. M. Smith et al. 2004). First, FA images were created by fitting a tensor model to the raw diffusion data using FDT, and then brain-extracted using BET (Smith 2002). FA data extracted from every DTI sequence from our cohort were then aligned into a common space using the nonlinear registration tool FNIRT (Andersson 2007a; Andersson 2007b), which uses a b-spline representation of the registration warp field (Rueckert et al. 1999). Next, the mean FA image was created and thinned to create a mean FA skeleton which represents the centers of all tracts common to the group. Each subject’s aligned FA data was then projected onto this skeleton and the resulting data fed into voxel-wise cross-subject statistics.

### 2.7 Computing Structural Connectivity from DTI

T1-weighted anatomical images were semi-automatically skull-stripped with BrainSuite’s Brain Extraction Sequence and automatically co-registered to the CONN atlas with BrainSuite’s surface volume registration (Shattuck and Leahy 2002). Using the BrainSuite Diffusion Pipeline, FA maps were derived from the DTI images and were co-registered with the T1-weighted anatomical image for each subject. Using BrainSuite’s diffusion toolbox, SC between the 132 ROIs was found based upon the DTI deterministic local tractography with a step-size of 0.25 mm using angular and FA thresholds of 35 degrees and 0.2, respectively (Hu et al. 2012; Ni et al. 2011; Min et al. 2014). The maximum number of steps was 500. The FA threshold ensures the tractography is based on white matter tracts which have high FA values as opposed to gray matter. The stop angle prevents the generation of fiber tracts that have a bend less than 35 degrees between any two consecutive points. The strength for a given structural connection was denoted by the number of fiber tracts that connects a particular node to another. Connectivity matrices containing the number of fiber tracts between each node-to-node connection were exported from BrainSuite (Shattuck et al. 2013).

### 2.8 Network Connectivity Group Comparison

In order to compare differences in structural and functional connectivity between mTBI and HC, we performed unthresholded independent two-sample t-tests on the connectivity matrices between mTBI and control groups using the SciPy python library (Jones, Oliphant, and Peterson 2001). In section 3.2, we show the t-scores from the comparison between mTBI and HC of each connection in both functional and structural connectomes. When performing feature selection for machine learning analysis, we employed the same method to compare groups but only compared subjects within a given training fold (see section 2.8).

### 2.9 Machine Learning Classification

In order to differentiate between mTBI and HC brain networks, we employed linear Support Vector Machine (SVM) models trained on the connectivity matrices from groups. We validated our model using a leave-one-out cross-validation (LOOCV) approach, where we repeatedly trained the SVM on all but one held-out subject. The predictions of the held-out subject data were then concatenated and compared to the true class labels. In order to maximize classification performance, we used statistical group comparisons to select a subset of network connections for classification within each cross-validation training fold. The entire analysis was repeated 20 times in order to verify the stability of predictions. All analysis was written in Python using the Sci-Kit Learn library (Rossum and Drake 1995; Pedregosa et al. 2011).

### 2.10 Selecting Network Connections for Classification

As shown in Figure 3, we first separately performed t-tests on structural and functional connectivity matrices between each group for all samples within a given training data set. We then measured the t-score of the group difference for each network connection. Next, we separately ranked functional and structural network connections according to their t-score. We then performed stepwise feature selection on the training data set, measuring the classification performance of the SVM trained on the network connections with the top N t-scores—where N increased between 1 and 20. When selecting features to classify the single held-out test sample, we selected the N connections that yielded the highest AUC in the training fold.

**Figure 2:**
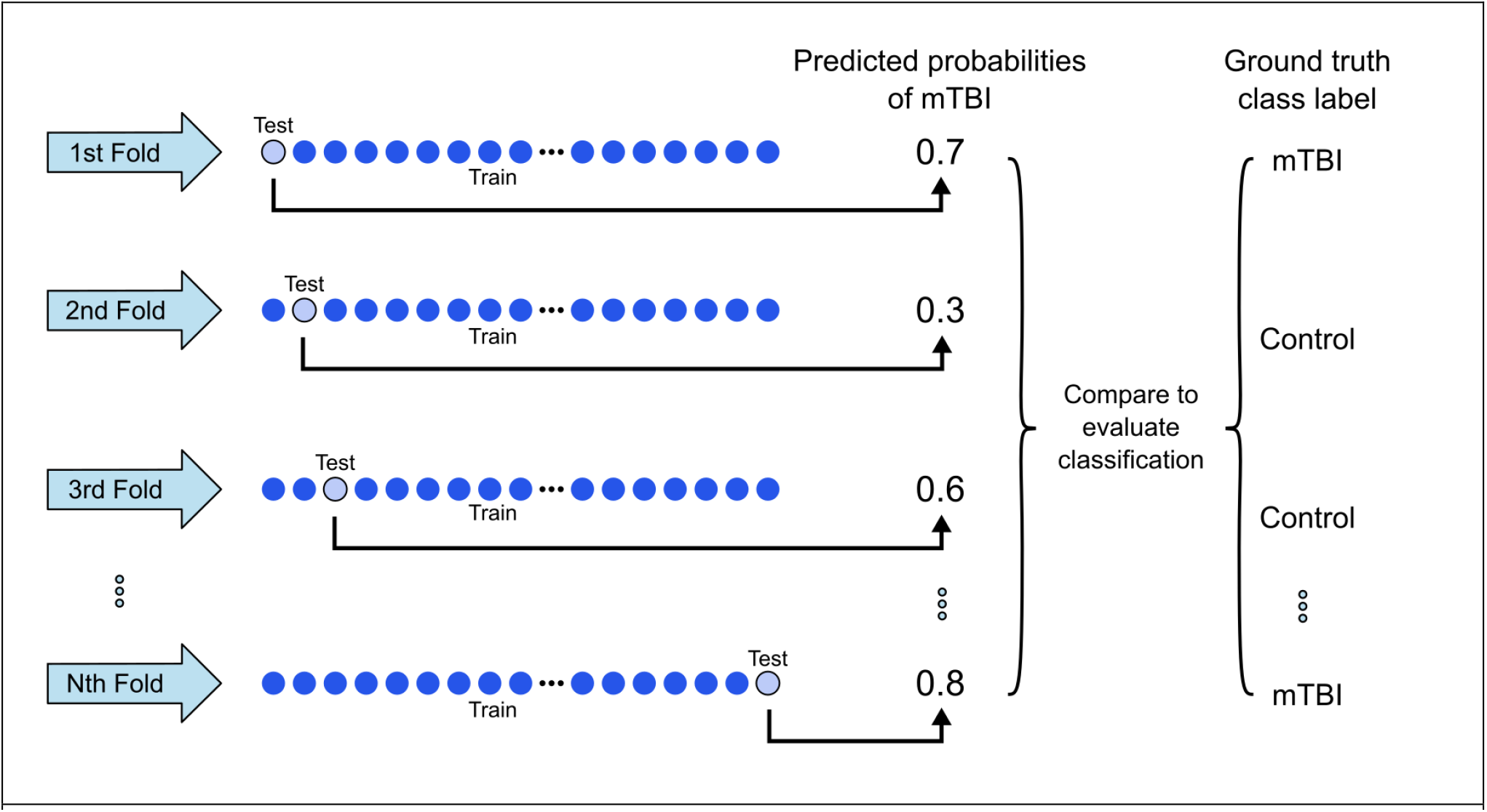
Leave-one-out cross validation procedure (LOOCV). Every test split (or “fold”) contains a single subject. The support vector machine (SVM) classifier is trained on all other subjects, and then the classifier predicts the probability of mTBI for the single held-out test sample. The data is repeatedly split so that every subject is used as a held-out test sample. Once the probability of mTBI has been predicted for all test samples, a probability threshold is selected such that test samples with predicted probabilities above the threshold are classified as mTBI and those with probabilities below the threshold are classified as HC. These classifications are compared to the ground truth class labels for each test sample, and the probability threshold is adjusted such that classification accuracy is maximized and sensitivity is non-zero. Figure adapted from (Shalbaf et al. 2020).

**Figure 3:**
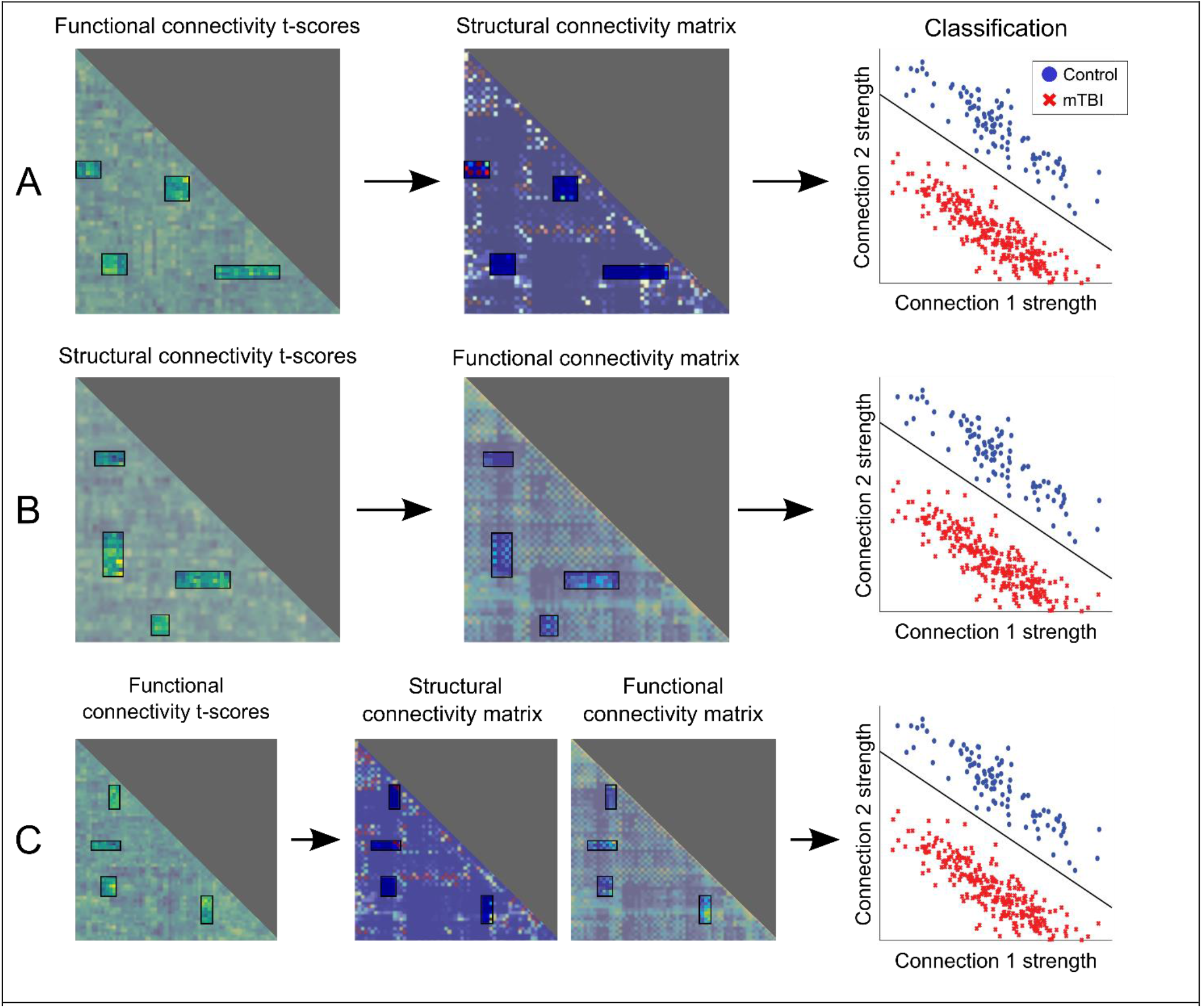
Here we depict our feature selection method for classification. Within each training fold, we performed group comparisons on either functional or structural connectivity matrices. We then identified the connections with the highest t-scores. Next, we selected those connections from either the structural or functional connectivity matrices—or both matrices—for classification. For example, in **A**, we performed a group comparison of functional connectivity matrices between mTBI and Control subjects, identified the functional connections with the highest t-scores, and then trained an SVM classifier on those respective structural connections to discriminate between mTBI and Control subjects. Artificially generated data is shown in the classification figures to demonstrate our methodology.

In total, there were 6 combinations of feature selection methods. They are listed as follows: (1) perform a t-test on functional connectivity matrices, identify N connections with the highest t-scores, and select those corresponding functional connections for classification. (2) perform a t-test on functional connectivity matrices, identify N connections with the highest t-scores, and select those corresponding structural connections for classification. (3) perform a t-test on functional connectivity matrices, identify N connections with the highest t-scores, and select those corresponding functional and structural connections for classification. (4) perform a t-test on structural connectivity matrices, identify N connections with the highest t-scores, and select those corresponding functional connections for classification. (5) perform a t-test on structural connectivity matrices, identify N connections with the highest t-scores, and select those corresponding structural connections for classification. (6) perform a t-test on structural connectivity matrices, identify N connections with the highest t-scores, and select those corresponding functional and structural connections for classification.

After classification analysis, we identified the 10 connections that were most frequently selected by the feature selection method that yielded the highest classification accuracy. We counted the number of times each of the 10 connections were selected. Next, we plotted the connections using the Nibabel 3.2.1 python package (Gramfort et al. 2014).

### 2.11 Performance Evaluation

For each LOOCV split, we recorded the predicted probability of mTBI for the held-out test sample. Next, we concatenated the 36 probabilities and used them to generate a ROC curve and evaluated the AUC. We then identified the optimal probability threshold that yielded the best accuracy and non-zero sensitivity. Among the predicted probabilities for the 37 test-samples, we classified probabilities above the selected threshold as “mTBI” and probabilities below this threshold as “Control”. Next, we compared these classifications to the actual class labels and reported the accuracy, sensitivity, specificity, and F1-score. After evaluation, we compared the AUC metrics of the best and second-best performing feature selection methods across repeated instantiations using an independent two-tailed t-test implemented using the SciPy library (Jones, Oliphant, and Peterson 2001).

### 2.12 DTI Classification

After performing feature selection on the structural and functional connectivity matrices, we determined the ROIs that participated in the most predictive connections. We then classified FA voxels within those ROIs. We classified both FA voxels as well as the mean FA values within a given ROI.

## 3 Results

### 3.1 Subject Data

Sixty-seven adult subjects (>18 years of age) each had MR exams acquired at our institution with the same acquisition protocol (Table 1). The MR acquisition protocol included rs-fMRI, DTI, and high-resolution non-contrast T1 anatomical image. Subjects whose ages were greater than 2.5 standard deviations from the mean group age or whose images were distorted by motion artifacts were excluded from our analysis. Twenty healthy subjects from our control database served as the control group and sixteen subjects with mTBI formed the pathological group. Demographics collected on eligible subjects included age and gender. The control group’s age was 26.5 ± 4.15 years (range: 22-40 years) and included 12 males and 8 females. The mTBI group’s age was 43.65 ± 8.37 years (range: 27-61 years) and included 10 male and 6 females. Statistical comparisons revealed that the mTBI and control groups differed in age (p<0.001) but not in sex (p=0.88).

**Table 1:** Patient demographics across controls and subjects with mild traumatic brain injury (mTBI) [N=36].

3.1 Subject Data
|  |  |  |  |
| --- | --- | --- | --- |
| Age (in years) | Control (n=20) | mTBI (n=16) | p-value |
| Mean ± SD | 26.5 ± 4.15 | 43.65 ± 8.37 | < 0.001 |
| 95% CI for Mean | 24.51–28.50 | 39.21–48.08 |  |
| Median (IQR) | 26.0 (4.5) | 46.0 (12.0) |  |
| Sex, n(%) |  |  | 0.88 |
| Male | 12 (60.0%) | 10 (62.5%) |  |
| Female | 8 (40.0%) | 6 (37.5%) |  |

### 3.2 Structural and Functional Connectivity Group Comparisons

Before performing machine learning analysis, we compared the functional and structural connectomes of subjects with mTBI to healthy controls to find connections that differed between the two groups.

Nine of the ten functional connections with the highest magnitude t-scores were decreased in connectivity in mTBI compared to control. These connections existed between areas in the frontal lobe, temporal lobe, and cerebellum (Table 2). In contrast, all ten structural connections with the highest t-scores were increased in connectivity in mTBI compared to control and existed between areas in the frontal and temporal lobes (Table 2).

**Table 2:**
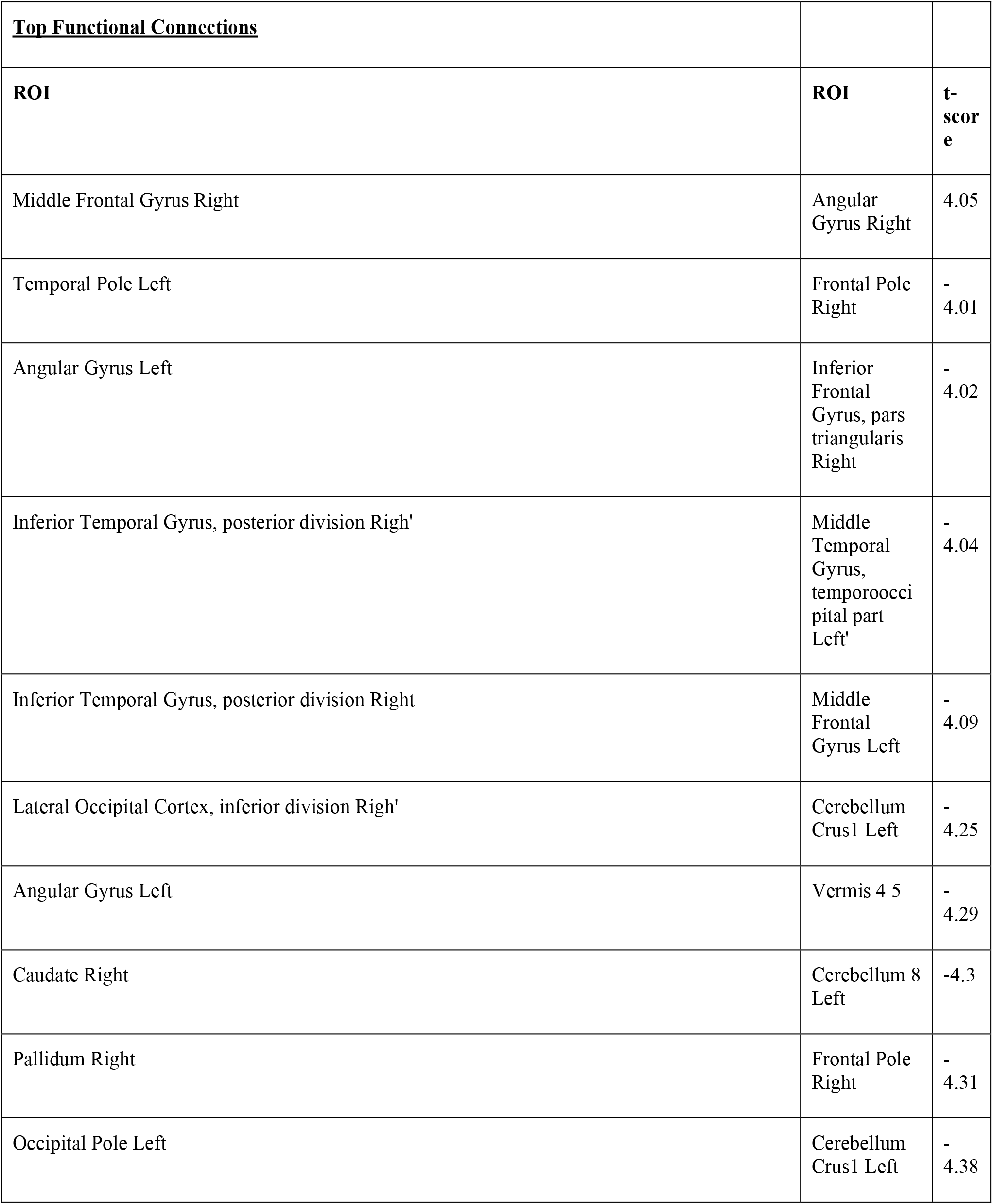
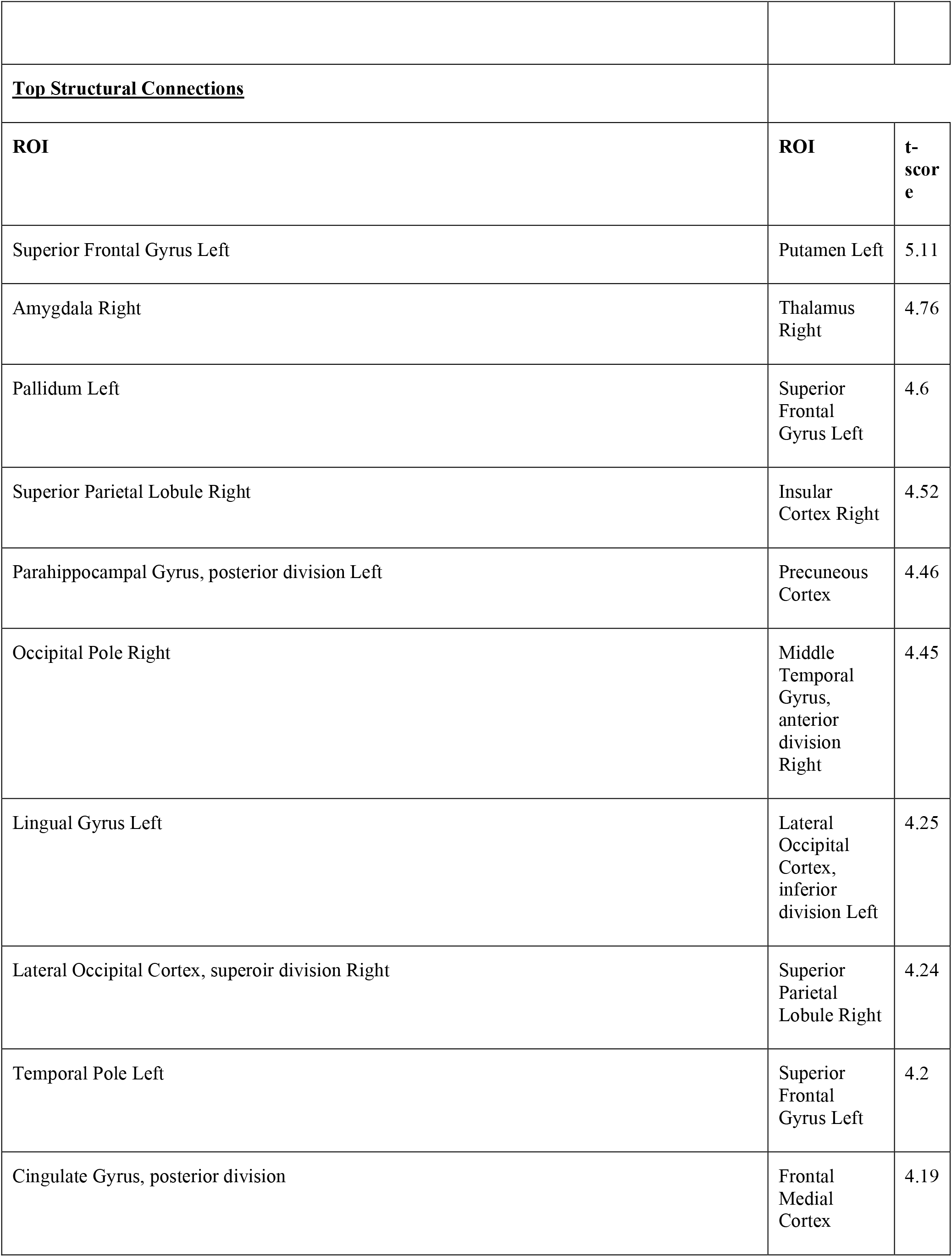
Connections with 10 highest t-scores.

|  |  |  |
| --- | --- | --- |
| connectivity (**B**). In each subplot, the identified connections were viewed in coronal, sagittal and axial projections from left to right. |  |  |
| **Table 2: Connections with 10 highest t-scores** |  |  |
| **<u>Top Functional Connections</u>** |  |  |
| **ROI** | **ROI** | **t-score** |
| Middle Frontal Gyrus Right | Angular Gyrus Right | 4.05 |
| Temporal Pole Left | Frontal Pole Right | - 4.01 |
| Angular Gyrus Left | Inferior Frontal Gyrus, pars triangularis Right | - 4.02 |
| Inferior Temporal Gyrus, posterior division Righ' | Middle Temporal Gyrus, temporooccipital part Left' | - 4.04 |
| Inferior Temporal Gyrus, posterior division Right | Middle Frontal Gyrus Left | - 4.09 |
| Lateral Occipital Cortex, inferior division Righ' | Cerebellum Crus1 Left | - 4.25 |
| Angular Gyrus Left | Vermis 4 5 | - 4.29 |
| Caudate Right | Cerebellum 8 Left | -4.3 |
| Pallidum Right | Frontal Pole Right | - 4.31 |
| Occipital Pole Left | Cerebellum Crus1 Left | - 4.38 |
| **<u>Top Structural Connections</u>** |  |  |
| **ROI** | **ROI** | **t-score** |
| Superior Frontal Gyrus Left | Putamen Left | 5.11 |
| Amygdala Right | Thalamus Right | 4.76 |
| Pallidum Left | Superior Frontal Gyrus Left | 4.6 |
| Superior Parietal Lobule Right | Insular Cortex Right | 4.52 |
| Parahippocampal Gyrus, posterior division Left | Precuneous Cortex | 4.46 |
| Occipital Pole Right | Middle Temporal Gyrus, anterior division Right | 4.45 |
| Lingual Gyrus Left | Lateral Occipital Cortex, inferior division Left | 4.25 |
| Lateral Occipital Cortex, superioir division Right | Superior Parietal Lobule Right | 4.24 |
| Temporal Pole Left | Superior Frontal Gyrus Left | 4.2 |
| Cingulate Gyrus, posterior division | Frontal Medial Cortex | 4.19 |

### 3.3 Functional differences indicate structural alterations in mild TBI

Next, we sought to determine which subset of structural and/or functional connections are most predictive of mild TBI. To this end, we trained separate linear support vector machines on subsets of structural and functional connections and determined the model’s performance. We selected subsets of connections based on the functional and structural connectivity group comparisons between mTBI and control subjects. Within each training-set, we identified connections with the highest t-score for each modality (fMRI and DTI), and then classified using the connectivity values of these connections for each imaging modality separately as well as together.

When classifying with structural connections (Figure 5), we found classification performance was highest when structural connections were selected according to the functional connectivity group comparison (Table 3). Classification performance decreased when classifying functional connections selected according to the functional connectivity group comparison. Classifying both structural and functional connections that were selected according to the functional group comparison yielded a similar classification performance as when classifying functional connections alone.

**Table 3:**
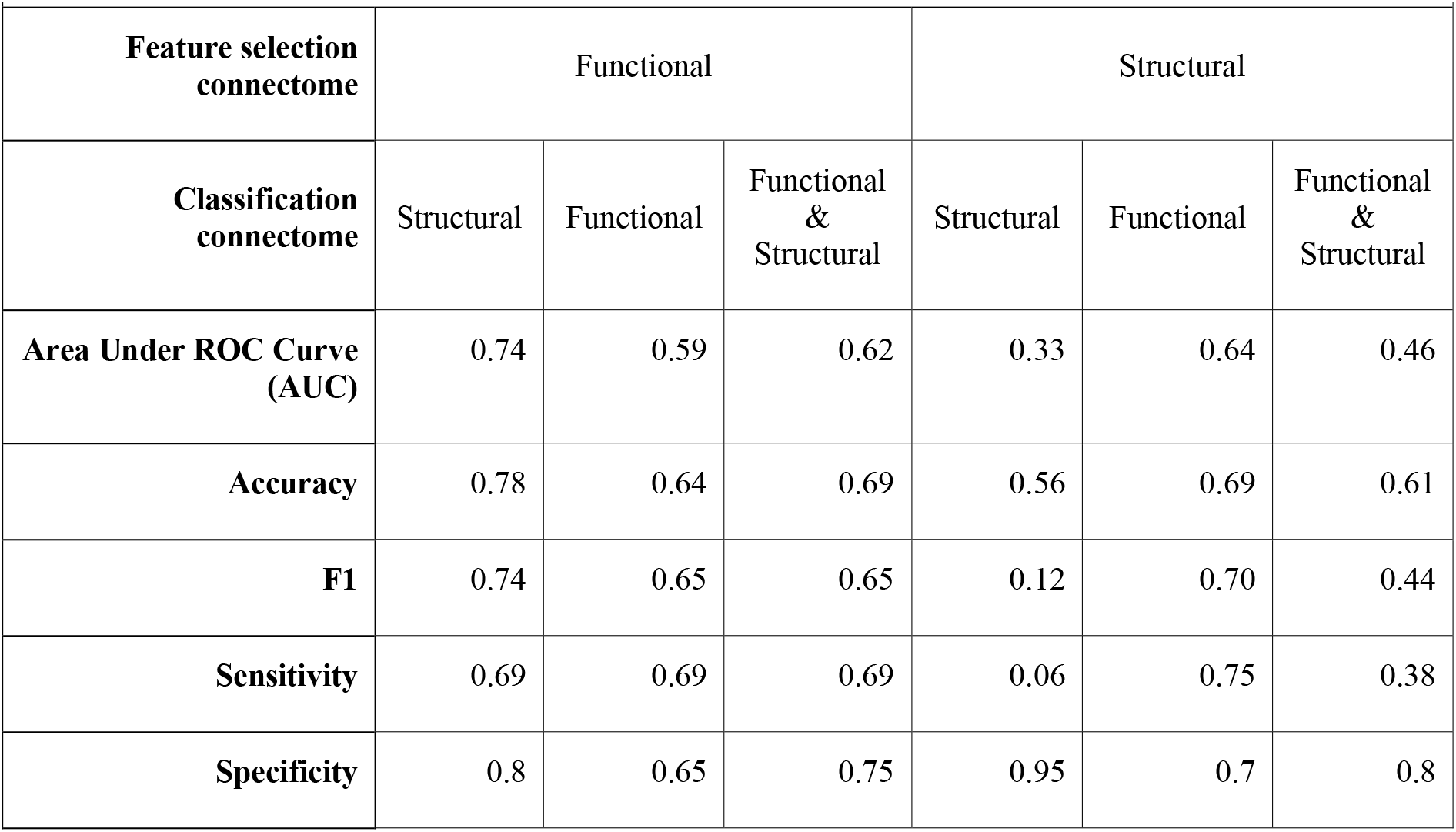
Performance metrics of different feature selection methods.

| Table 3: Performance metrics of different feature selection methods |  |  |  |  |  |  |
| --- | --- | --- | --- | --- | --- | --- |
| Feature selection connectome | Functional |  |  | Structural |  |  |
| Classification connectome | Structural | Functional | Functional & Structural | Structural | Functional | Functional & Structural |
| Area Under ROC Curve (AUC) | 0.74 | 0.59 | 0.62 | 0.33 | 0.64 | 0.46 |
| Accuracy | 0.78 | 0.64 | 0.69 | 0.56 | 0.69 | 0.61 |
| F1 | 0.74 | 0.65 | 0.65 | 0.12 | 0.70 | 0.44 |
| Sensitivity | 0.69 | 0.69 | 0.69 | 0.06 | 0.75 | 0.38 |
| Specificity | 0.8 | 0.65 | 0.75 | 0.95 | 0.7 | 0.8 |

**Table 3:** T-scores of most predictive functional connections identified by classification.

| ROI |  |  |
| --- | --- | --- |
|  | ROI | t-score |
| Middle Frontal Gyrus Right |  |  |
| Inferior Temporal Gyrus, posterior division Right | Angular Gyrus Right | 4.05 |
| Lateral Occipital Cortex, inferior division Right | Middle Temporal Gyrus, temporooccipital part Left | -4.01 |
| Angular Gyrus Left | Cerebellum Crus1 Left | - 4.02 |
| Angular Gyrus Left | Vermis 4 5 | - 4.04 |
| Pallidum Right | Inferior Frontal Gyrus, pars triangularis Right | - 4.09 |
| Caudate Right | Frontal Pole Right | - 4.25 |
| Middle Frontal Gryus Left | Cerebellum 8 Left | - 4.29 |
| Supramarginal Gyrus, anterior division Left | Pallidum Left | -4.3 |
| 'Temporal Pole Left' | Lateral Occipital Cortex, inferior division Left | - 4.31 |

**Figure 4:**
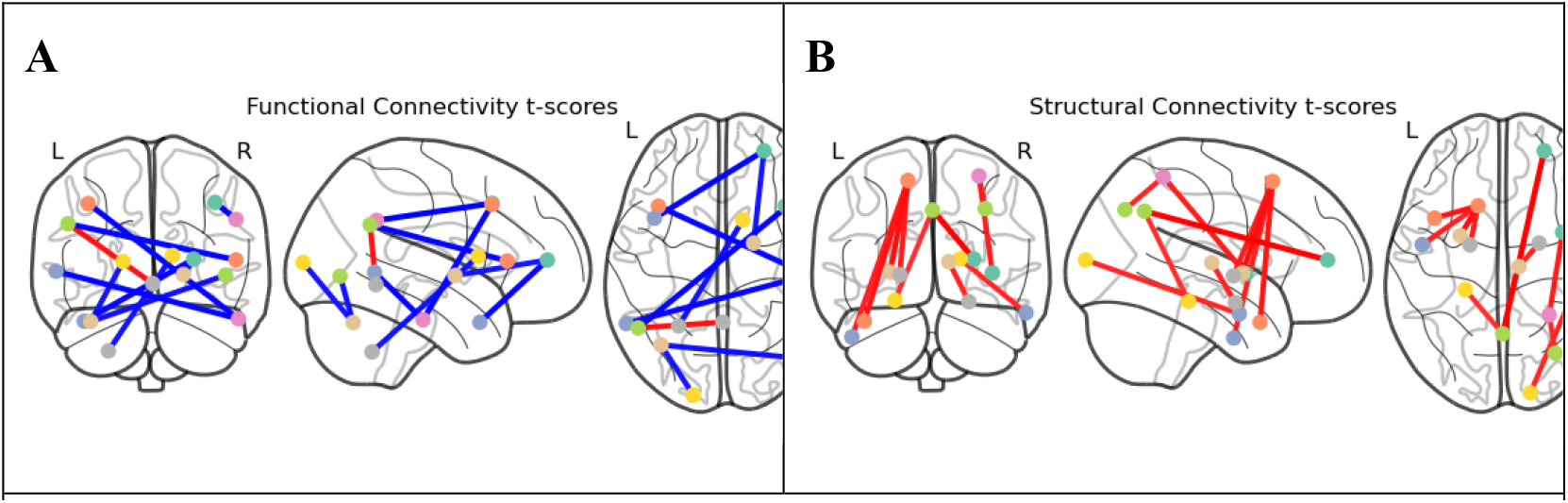
**S**hown here are the top 10 connections with highest absolute differences between mTBI and control groups estimated via two-tailed t-tests in both functional connectivity (**A**) and structural connectivity (**B**). In each subplot, the identified connections were viewed in coronal, sagittal and axial projections from left to right.

**Figure 5:**
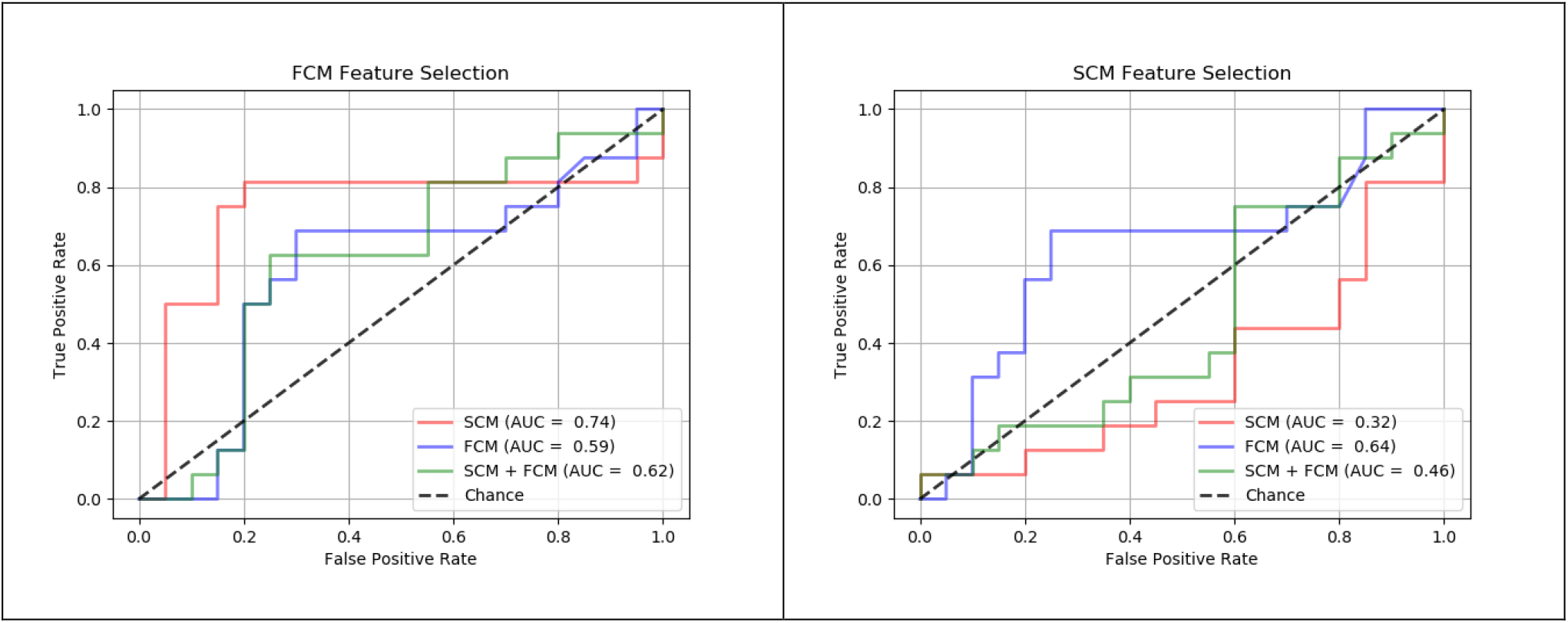
Here, we show the ROC curves for SVM classifiers trained according to the six different feature selection methods. We found that classifying the structural connections whose respective functional connections were the most different between mTBI and control yielded the highest classification performance.

Conversely, we found that classification performance was higher when classifying functional connections compared to classifying structural connections identified from the structural connectivity group comparison (Table 3). Classifying both structural and functional connections yielded a similar performance as when classifying with functional connectivity alone.

In order to determine if the differences in classification accuracy imparted by each feature selection method were stable across different random SVM initializations, we repeated each classification strategy 20 times. In Figure 3, we show the distribution of classification accuracies measured by AUC. Our results suggest that classifying structural connections selected by functional connectivity group differences consistently resulted in a higher classification accuracy. In comparison to the second-best performing feature selection method, classification of structural connections selected by functional connectivity group differences yielded a significantly higher AUC (p<0.05).

### 3.4 Mild TBI results in altered connectivity in the frontal and temporal lobes and cerebellum

During classification, we counted the number of times each connection was selected for classification during our stepwise feature selection procedure. Next, we plotted the top 10 most frequently selected connections. Many of the ROIs that participated in these top-10 connections were in the frontal and temporal lobes. Additionally, ROIs were in the Angular Gyrus, Supplementary Motor Cortex, and Cerebellum (Table 2).

### 3.5 FA Classification

We also compared classification performance of an SVM built on FA voxels and average FA values within the ROIs of the top 5 most frequently selected brain connections for SCM classification (Table 3). First, we performed an independent 2-tailed t-test of FA values between mTBI and controls and found no significant differences. Next, we found that classifying on FA values consistently yielded poor classification performance (Figure 8), which suggests that differences in brain structure after mTBI are more easily identified with large-scale measures such as tractography. In comparison to directly classifying FA voxels, classification of structural connections selected by functional connectivity group differences yielded a significantly higher AUC (p<0.001).

**Figure 6:**
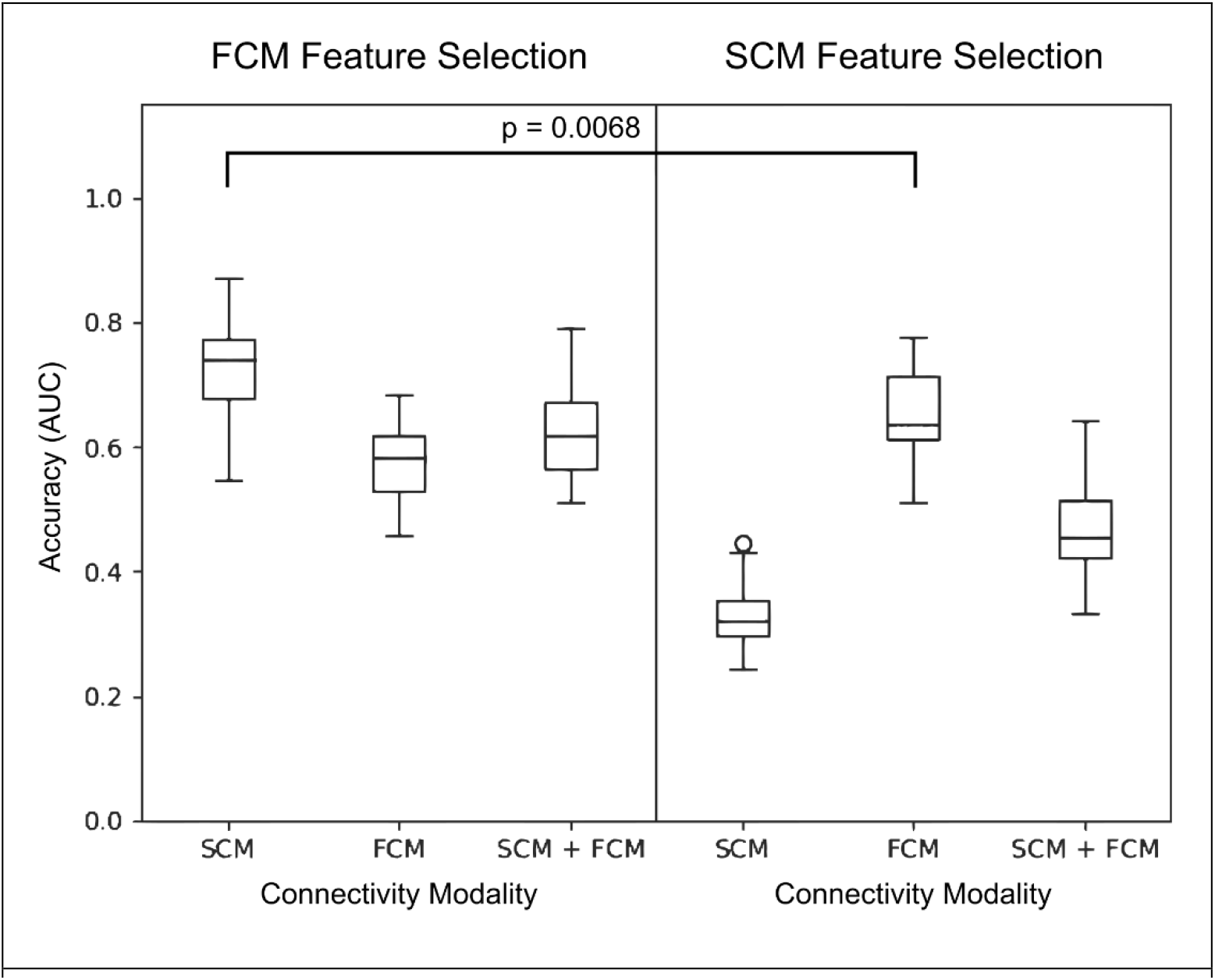
Shown are distributions of AUC accuracy values for SVM classifiers trained according to each of the six feature selection methods. In order to determine whether the differences in classification performance across the feature selection methods were due to random initializations of the SVM classifier, we repeated the entire training process 20 times. We found that selecting structural connections for classification based on the FCM group comparison yielded the highest classification accuracy measured by area under the ROC curve (AUC). * - statistical comparison where p < 0.05.

**Figure 7:**
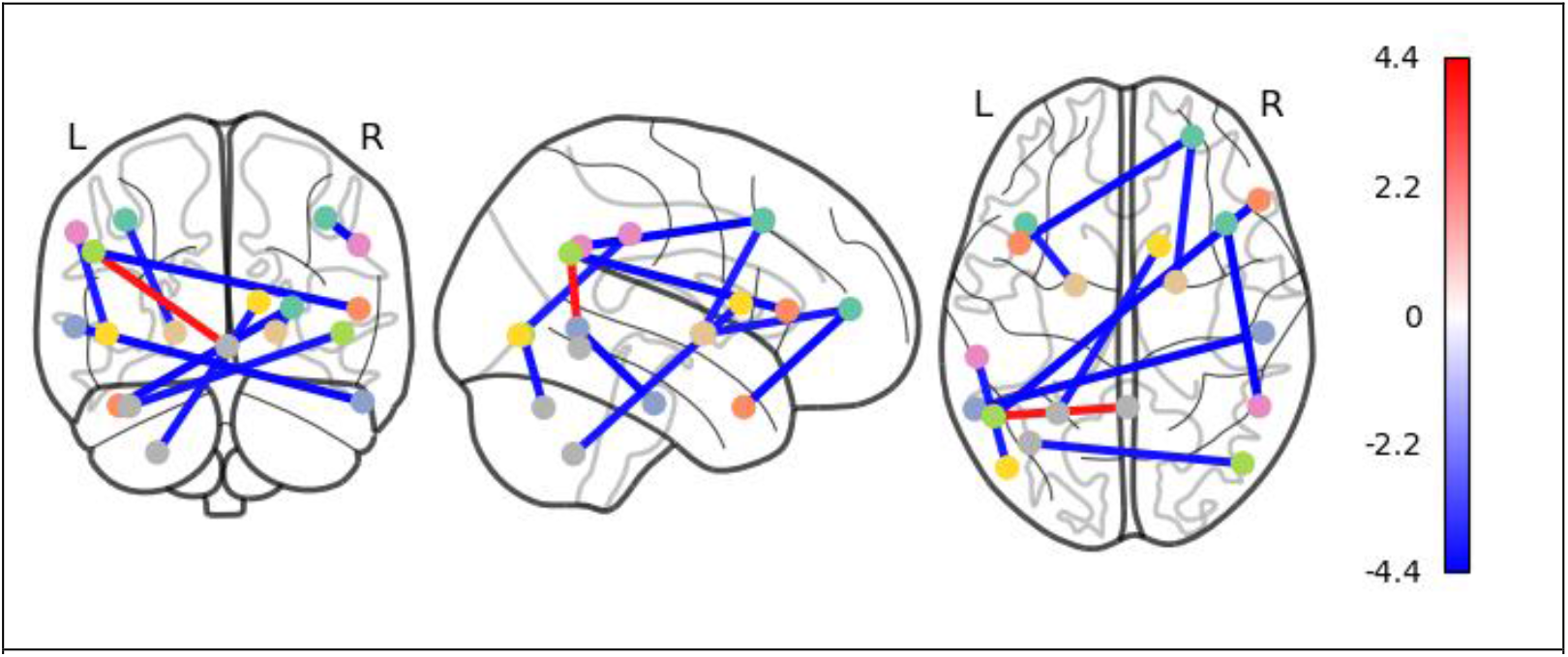
Here, we plotted the functional connectivity t-scores of the top 10 most frequently selected connections for classification. Discriminative connections were primarily between areas in the frontal lobe, temporal lobe, and cerebellum. In each subplot, the identified connections were viewed in coronal, sagittal and axial projections from left to right.

**Figure 8:**
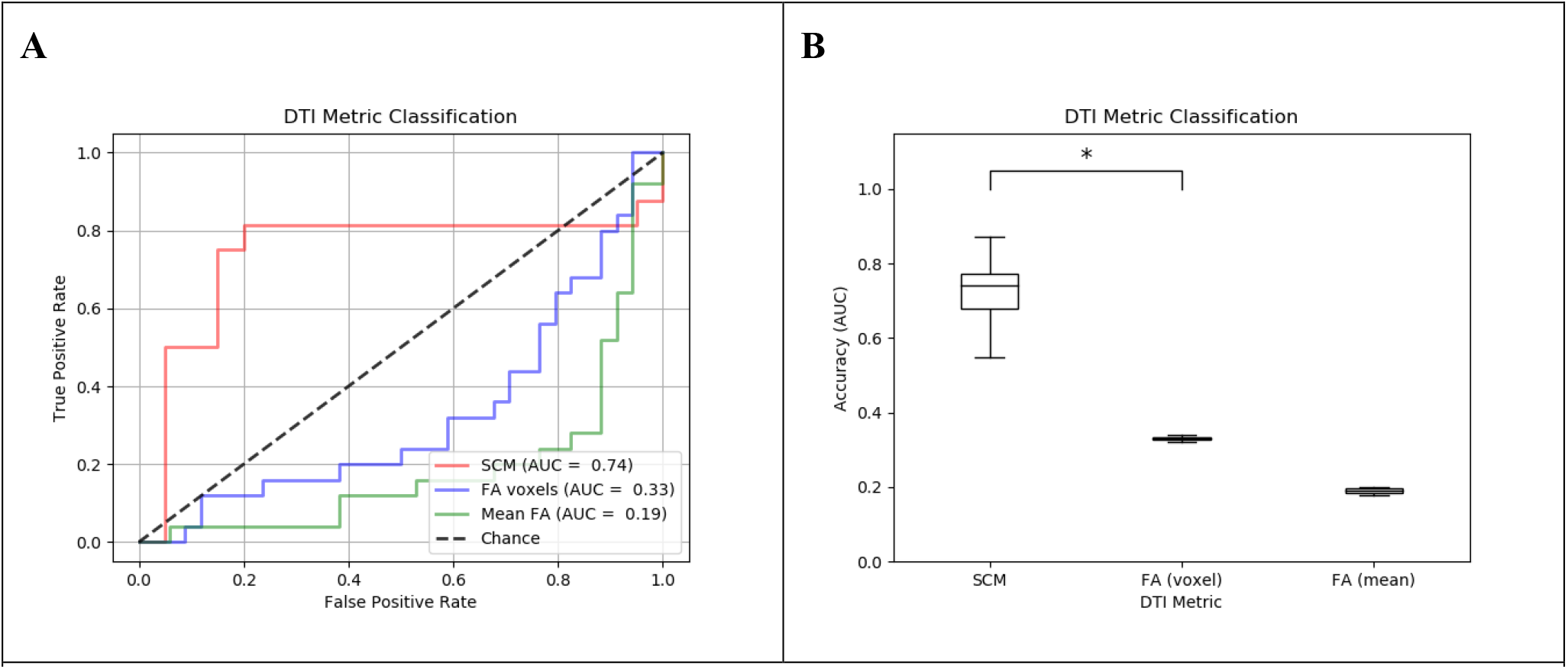
Here, we show the ROC curves and AUC accuracy metrics for SVM classifiers trained according to the best of the six feature selection methods as well as FA voxels and mean FA values. We determined whether structural connections, which are measured by diffusion tractography, are more predictive of mTBI than FA voxels or mean FA values within a given ROI. We created masks of the ROIs participating in the top 5 most frequently selected structural connections for classification (Table 3) and masked the FA data for all subjects. We compared the classification accuracy of the best performing feature selection method (classifying SCM selected according to FCM group comparison) to SVMs trained on all FA voxels in the selected ROIs as well as the average FA values of each ROI. In **A**, we show the ROC curves generated by the trained SVM classifiers, and in **B** we show the AUC performance metric measured across 20 different random initializations of SVM classifiers. We found that classifying an optimal subset of structural brain connections measured by diffusion tractography resulted in substantially better performance than classifying diffusions metrics. * - statistical comparison where p < 0.001.

## 4 Discussion

In this study, we applied a machine learning algorithm to classify subjects with mTBI from healthy controls using multimodal neuroimaging sequences and identified structural and functional networks that are altered in mTBI. Results from our functional connectivity analysis revealed a widespread hyperconnectivity and localized hypoconnectivity within the inferior temporal gyrus, brain stem, and cerebellum in mTBI subjects compared to healthy controls. Analysis of structural connectivity revealed widespread decreases and localized increases within the frontal and temporal cortical areas and several subcortical regions including the thalamus, cerebellum, and vermis.

Using a linear support vector machine validated by leave-one-out cross validation, we achieved a maximum classification accuracy of 78%. Our results show that structural connections across the frontal lobe, temporal lobe, supplemental motor area, and cerebellum were the most discriminative of mTBI (Figure 7; Table 3). Interestingly, our feature selection method revealed that the structural connection regions whose respective functional connections exhibited the largest differences between mTBI and control were the most predictive of mTBI. (Figure 4; Table 2). This suggests that changes in structural connectivity caused by mTBI may be identified by differences in functional connectivity.

### 4.1 Comparison to Current Literature

Although functional connectivity was increased in mTBI patients compared to control as demonstrated by qualitative inspection, the functional connections with the largest differences were decreased in mTBI. While past literature suggests that mTBI results in functional hyperconnectivity (Hayes, Bigler, and Verfaellie 2016), several studies have suggested that the direction of connectivity changes may be dependent on the phase of recovery from mTBI, and that functional hypoconnectivity is an early response to injury (Dall’Acqua et al. 2017; Zhu et al. 2015; Iraji et al. 2015).

Conversely, we found that the largest differences in structural connectivity were positive, while most other differences in structural connectivity were negative. Increases in structural connectivity among a small subset of network connections has been attributed to the “rich-club” hypothesis, whereby injury to tertiary nodes triggers a rerouting of neural architecture towards more central network hubs (Dall’Acqua et al. 2017; Heuvel and Sporns 2011)

(Mitra et al. 2016)Previous studies have analyzed unimodal neuroimaging datasets to discriminate mTBI from healthy control. A study that attempted classification on unimodal datasets to detect mTBI via structural connections was by (Mitra et al. 2016). The authors achieved a classification accuracy of 68%. In contrast, our method demonstrated that multimodal imaging increased classification accuracy up to 78%.

Previous studies have shown that classification analysis using multimodal datasets either decrease classification performance (Vergara et al., 2016), or cannot identify a specific set of network connections predictive of mTBI (Sinke et al, 2021). However, our approach demonstrates that DTI and fMRI can be combined to yield high classification performance. Specifically, we showed that group differences in functional connectivity could be used to identify structural features predictive of mTBI. Our high classification performance was enabled by our novel multimodal feature selection method, which reduced the number of structural connections for classification.

An important component of our feature selection method was the identification of group differences in functional connectivity. It is well known that mTBI affects resting state functional connectivity and has been reported frequently in past literature (Mayer et al. 2011) (Stevens et al. 2012) (Palacios et al. 2017) (Iraji et al. 2015).

In comparison to fractional anisotropy (FA), we found that structural connectivity measured by tractography is more predictive of mTBI. Interestingly, this suggests that despite mTBI being normally associated with the cellular-level damage, metrics that capture fine-grain differences in white matter integrity are less predictive of mTBI. Instead, coarse-grained diffusion tractography captured network-level reorganization in patients with mTBI. This suggests that network-level analysis may yield more sensitive biomarkers to mTBI in comparison to diffusion tensor metrics. Additionally, our corroborate past results from Iraji et al who showed that traumatic brain injury results in connectome-scale reorganization (Iraji et al. 2016b). Finally, FA can be distorted by edema or other microenvironmental changes that are unrelated to white matter structural integrity.

### 4.2 Structural and Functional Connectivity Alterations in mTBI

Our multimodal approach using both DTI and rs-fMRI allowed us to assess the relationship between alterations in structural and functional connectivity in mTBI (Hayes, 2016). Specifically, we found that network connections with altered functional connectivity also exhibit structural connectivity that is predictive of mTBI. Relationships between structural and functional connectivity have been identified in past literature and are termed “structure-function coupling” (Honey et al. 2009). In mTBI, disruption or “decoupling” between structural and functional connectomes has been associated with clinically relevant symptoms such as cognitive sensorimotor and behavioral impairments (Harris, Verley, Gutman, Thompson, et al., 2016; Sinke et al., 2021).

The literature has reported varying effects of mTBI on structural-functional coupling. Several studies have found positive correlations between changes in structural and functional connectivity in mTBI (Palacios et al. 2013b) (Zhang et al. 2010) (Sharp et al. 2011). Other studies have either found that both structural and functional connectivity is decreased or that an inverse relationship between the two connectomes occurs in mTBI. Wang et al found a negative correlation between structural and functional connectivity in mTBI (Wang et al. 2021)Rajesh et al found mTBI was associated with functional hypo-activation across the default mode network, which corroborates the decrease in functional connectivity among the most predictive connections in our study (Rajesh et al. 2017). Tang et al also identified an inverse relationship between fractional anisotropy and default mode network functional connectivity (Tang et al. 2012).Other studies have either found that both structural and functional connectivity is decreased or that mTBI exhibits an inverse relationship between the two connectomes. (Wang et al. 2021)(Rajesh et al. 2017) (Tang et al. 2012)

In contrast, our results indicate a more local relationship between structural and functional connectivity, whereby physical injury to structural connections between the frontal lobe and temporal lobes results in altered functional connectivity among those same connections. Our findings have been corroborated by Iraji et al, who identified the same “landmark” structural connections that also exhibited functional alterations (Iraji et al. 2016b). Other studies have also reported local differences in functional and structural connectivity in mild TBI (Harris, 2016) (Sharp, 2011).

We found that structural connections across the frontal and temporal lobes were predictive of mTBI. Interestingly, both the temporal and frontal lobes are particularly vulnerable to injury due to its sensitivity to inertial forces, which has been demonstrated in numerous human and animal studies (Bigler 2007; Kampfl et al. 1998; Kotapka et al. 1991; Smith et al. 1997; Cullen et al. 2016)

A possible mechanism behind the alterations to structure-function coupling after mTBI has been proposed by Kuceyeski et al in 2019. The authors suggest that increased structural and functional network changes are a compensatory response to mTBI (Kuceyeski et al. 2019). Future studies can explore the mechanisms underlying network reorganization after mild TBI.

## 5 Limitations and Future Directions

Our study has several limitations. First, subjects with TBI can suffer injury to any nonspecific brain area. This heterogeneity presents a challenge in generalizing our results to the larger patient population. Related to this issue is the small sample size present in our study. Evidence has shown that performance of machine learning algorithms scales with sample size in neuroimaging studies (Marc-Andre et al. 2020). A separate limitation is that the age of our mTBI cohort is older than the control group. Additionally, our study did not take into consideration the heterogeneity of the timing of neuroimaging relative to the timing of injury among our subjects. The phase of recovery after the brain injury has been shown to have an impact on changes in brain connectivity (Dall’Acqua et al. 2017). In order to overcome these limitations, longitudinal and prospective studies are needed to gain a better understanding of how early connectivity changes due to mTBI result in chronic cognitive and behavioral deficits. Better study design may reveal more sensitive and specific biomarkers that can be used in tandem with clinical evaluation to enable early diagnosis. In the future, longitudinal studies are needed to gain a better understanding of how early connectivity changes due to mTBI result in chronic cognitive and behavioral deficits. Prospective studies may reveal specific biomarkers that can be used in tandem with clinical evaluation to enable early diagnosis.

## 6 Conclusion

In summary, our machine-learning approach revealed changes in brain networks associated with mild TBI. Through a novel feature selection method, we demonstrated that multimodal imaging of both structural and functional connectomes can serve as a potential biomarker for mild TBI. Our results show that differences in functional connectivity are reflected by corresponding changes in structural connectivity. Further, our results suggest that mild TBI affects the relationship between structural and functional connectivity.

## Data Availability

All data produced in the present study are available upon reasonable request to the authors

## 8 Author Contributions

Conceptualization, A.P. and H.S.; methodology, A.P., H.S., K.H., and B.J.B.; image processing: K.H., B.J.B., D.M., R.D., P.H., T.P., A.P., and T.S.; software, A.P. and K.H.; formal analysis, A.P.; investigation, A.P., T.S, H.S. C.L., and B.B.; administrative support and resources, H.S., C.L., E.G., C.T. and T.Q.; data curation, C.L., D.V., E.G., T.Q., and R.R., writing—original draft preparation, A.P., T.S., K.H. and C.T.; writing—review and editing, A.P., T.S, P.K., C.L., D.V., B. Biswal, and H.S.; visualization, C.T. and A.P.; supervision, H.S. and B. Biswal.

## 10 Acknowledgments

The authors would like to acknowledge the support from Department of Neurosurgery and Department of Radiology at Louisiana State University Health Sciences Center in Shreveport, LA.

## 11 Data Availability Statement

Data and code for this study will be made available upon request.

